# Predictors of COVID-19 vaccine hesitancy in the UK Household Longitudinal Study

**DOI:** 10.1101/2020.12.27.20248899

**Authors:** Elaine Robertson, Kelly S Reeve, Claire L Niedzwiedz, Jamie Moore, Margaret Blake, Michael Green, Srinivasa Vittal Katikireddi, Michaela J Benzeval

## Abstract

**Background:** Vaccination is crucial to address the COVID-19 pandemic but vaccine hesitancy could undermine control efforts. We aimed to investigate the prevalence of COVID-19 vaccine hesitancy in the UK population, identify which population subgroups are more likely to be vaccine hesitant, and report stated reasons for vaccine hesitancy.

**Methods:** Nationally representative survey data from 12,035 participants were collected from 24th November to 1st December 2020 for wave 6 of the ‘Understanding Society’ COVID-19 web survey. Participants were asked how likely or unlikely they would be to have a vaccine if offered and their main reason for hesitancy. Cross-sectional analysis assessed prevalence of vaccine hesitancy and logistic regression models conducted.

**Findings:** Overall intention to be vaccinated was high (82% likely/very likely). Vaccine hesitancy was higher in women (21.0% vs 14.7%), younger age groups (26.5% in 16-24 year olds vs 4.5% in 75+) and less educated (18.6% no qualifications vs 13.2% degree qualified). Vaccine hesitancy was particularly high in Black (71.8%), Pakistani/Bangladeshi (42.3%), Mixed (32.4%) and non-UK/Irish White (26.4%) ethnic groups. Fully adjusted models showed gender, education and ethnicity were independently associated with vaccine hesitancy. Odds ratios for vaccine hesitancy were 12.96 (95% CI:7.34, 22.89) in the Black/Black British and 2.31 (95% CI:1.55, 3.44) in Pakistani/Bangladeshi ethnic groups (compared to White British/Irish ethnicity) and 3.24 (95%CI:1.93, 5.45) for people with no qualifications compared to degree educated. The main reason for hesitancy was fears over unknown future effects.

**Interpretation:** Older people at greatest COVID-19 mortality risk expressed the greatest willingness to be vaccinated but Black and Pakistani/Bangladeshi ethnic groups had greater vaccine hesitancy. Vaccine programmes should prioritise measures to improve uptake in specific minority ethnic groups.

**Funding:** Medical Research Council

**Research in context:** *Evidence before this study:* We searched Embase and Medline up to November 16, 2020, using key words “vaccine hesitancy” and “COVID-19” or “SARS-CoV-2”. Vaccine hesitancy is complex but also context specific. Previous research about vaccine hesitancy relates to existing adult and childhood vaccines, with limited evidence currently available on willingness to be vaccinated for newly available COVID-19 vaccines. Existing vaccination programmes often have lower uptake among more socioeconomically disadvantaged groups. Uptake of vaccines has often varied across ethnic groups, but patterns have often varied across different vaccine programmes.

*Added value of this study:* Our study describes the sub-groups of the UK population who are more likely to be hesitant about a COVID-19 vaccine and examines possible explanations for this. We used nationally representative data from the COVID-19 survey element of the UK’s largest household panel study. We asked specifically about vaccine hesitancy in relation to a COVID-19 vaccine at a time when initial results of vaccine trials were being reported in the media. We found willingness to be vaccinated is generally high across the UK population but marked differences exist across population subgroups. Willingness to be vaccinated was greater in older age groups and in men. However, some minority ethnic groups, particularly Black/Black British and Pakistani/Bangladeshi, had high levels of vaccine hesitancy but this was not seen across all minority ethnic groups. People with lower education levels were also more likely to be vaccine hesitant.

*Implications of all the available evidence:* The current evidence base on vaccine hesitancy in relation to COVID-19 is rapidly emerging but remains limited. Polling data has also found relatively high levels of willingness to take up a COVID-19 vaccine and suggested greater risks of vaccine hesitancy among Black, Asian and Minority Ethnic (BAME) people. Our study suggests that the risk of vaccine hesitancy differs across minority ethnic groups considerably, with Black ethnic groups particularly likely to be vaccine hesitant within the UK. Some White minority ethnic groups are also more likely to be vaccine hesitant than White British/Irish people. Herd immunity may be achievable through vaccination in the UK but a focus on specific ethnic minority and socioeconomic groups is needed to ensure an equitable vaccination programme.

## INTRODUCTION

Vaccines and immunisation programmes save lives. The World Health Organisation (WHO) estimates that immunisation programmes across the world prevent 2-3 million deaths every year and are not only cost effective but a key element of preventative healthcare.^1^ Since the emergence of the Severe Acute Respiratory Syndrome Coronavirus 2 (SARS-CoV-2) in 2019, the global pandemic has resulted in over 72 million confirmed cases and 1.6 million deaths in 220 countries worldwide (as at 18 Dec 2020).^2^ An effective and safe vaccine is vital to controlling the COVID-19 outbreak. However, not only does a vaccine need to be safe and effective, it must also be taken up by those people at greatest risk of harm from the disease. Ideally, uptake by a large enough proportion of the population will offer protection to people who remain unimmunised, referred to as achieving ‘herd immunity’. For COVID-19, vaccine uptake would need to be between approximately 67% and 80% to reduce spread of the disease.^3,4^ Understanding who will take up a vaccine, who plans not to or are uncertain, and why, is critical to designing a successful vaccination programme.

Even before the emergence of SARS-CoV-2, WHO had already highlighted vaccine hesitancy as one of the ten leading threats to global health.^5^ The WHO Strategic Advisory Group of Experts Working Group on Vaccine Hesitancy define it as the “*delay in acceptance or refusal of safe vaccines despite availability of vaccination services*”.^6^ Reasons for vaccine hesitancy are acknowledged as being complex but also context specific. Hesitancy around uptake can vary geographically, at different times and for different vaccines by a range of factors including complacency around the disease, convenience of access and confidence in the vaccine itself. For example, in the H1N1 pandemic of 2009, there was a perception that the vaccine was rushed and unsafe.^7^ Similar concerns have been raised about the speed of COVID-19 vaccine development.^4^

The COVID-19 pandemic has been described as a ‘syndemic’ where it not only acts together with, but worsens and amplifies, non-communicable diseases and social conditions and hence exacerbates inequalities in health.^8^ As inequalities already exist in seasonal influenza and pneumococcal vaccine uptake, with lower uptake in more deprived areas ^9,10^, there is reason to believe that inequalities will exist in vaccine uptake against COVID-19. Understanding if some subgroups of the population are more likely to be vaccine hesitant will help in the formulation of vaccination programme strategies to ensure adequate population coverage to achieve herd immunity and minimise health inequalities. This will be particularly important if greater vaccine hesitancy is found among groups at greater risk from COVID-19. We therefore aimed to describe how willing the UK population is to be vaccinated against COVID-19, to identify which population subgroups are more likely to be vaccine hesitant, which are more likely to take up a vaccine, and describe the main reasons given for both vaccine uptake and vaccine hesitancy.

## METHODS

### Data

The UK Household Longitudinal Study, also referred to as ‘Understanding Society’, is a nationally representative panel study, based on a clustered-stratified probability sample of UK households, with boost samples of key ethnic minority groups. Sample members living in the UK have been interviewed annually since 2009.^11^ In 2020, during the COVID-19 pandemic, participants aged 16+ years who had lived in households that had completed at least one of the last two waves of the main Understanding Society survey were invited to take part in the COVID-19 survey either online or by phone (n=42,330). Web surveys took place monthly from April to July, then every two months.^12^ For wave 6, carried out from 24^th^ November to 1^st^ December 2020, only sample members who had completed at least one partial interview in any of the preceding five COVID-19 web surveys, and had not become ineligible through death or moving abroad nor opted out of the study, were invited to take part. This resulted in 19,289 invitations being issued, and 12,035 took part, a response rate of 62%.^13^ Data for age, sex, ethnicity, education level and country of birth were derived from previous waves of the main study.^14^

Ethics approval was granted by the University of Essex Ethics Committee for the COVID-19 surveys (ETH1920-1271). No additional ethical approval was necessary for this secondary data analysis.

### Outcome Measures

Our primary outcome was vaccine hesitancy, assessed by asking all participants “Imagine that a vaccine against COVID-19 was available for anyone who wanted it. How likely or unlikely would you be to take the vaccine?” Possible responses were “Very likely”, “Likely”, “Unlikely” and “Very unlikely”. This was collapsed into a binary variable for modelling comparing ‘unlikely and very unlikely’, classified as vaccine hesitant, to ‘likely or very likely’. If a participant tried to bypass the question, they were given the option of ‘don’t know’(n=45) and, given the size of the group they were coded as missing.

Secondary outcomes included reasons for vaccine hesitancy. Vaccine hesitant participants were asked “What is the main reason you would not take the vaccine?” and asked to pick one main reason from a list of 12 possible answers (Figure 3). Participants who were not vaccine hesitant were asked “What would be your main reason for taking the vaccine?” and asked to choose one main reason from a list of 11 possible answers (Figure 3). All participants were then asked “*Which three of these things would most increase the chances of you choosing to get vaccinated*?” and given a list of 9 possible answers (Figure 3). For the purpose of this analysis, we identified if the item was mentioned as any of the three options.

**Figure 1.**
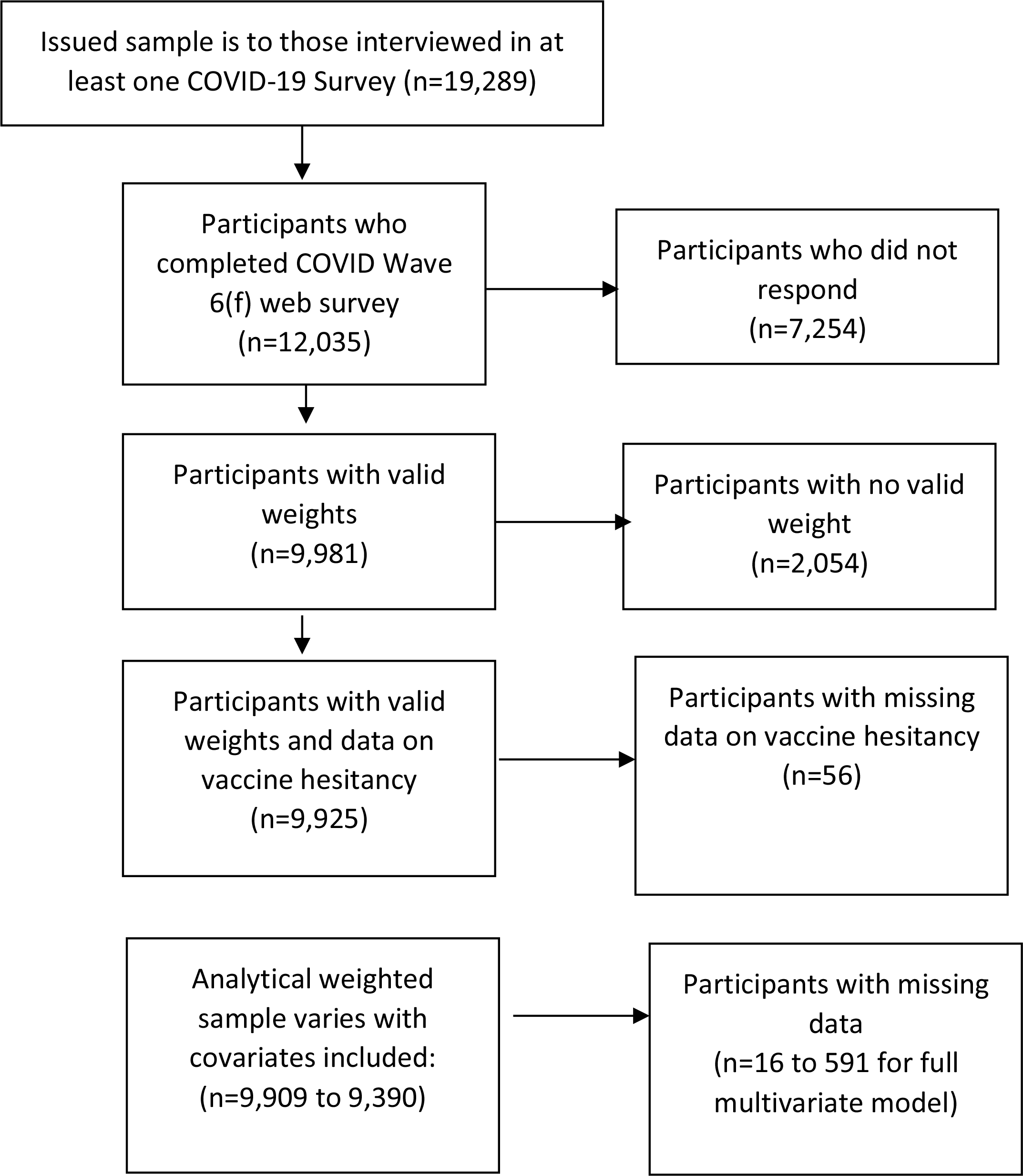
Selection of analytical sample.

**Figure 2.**
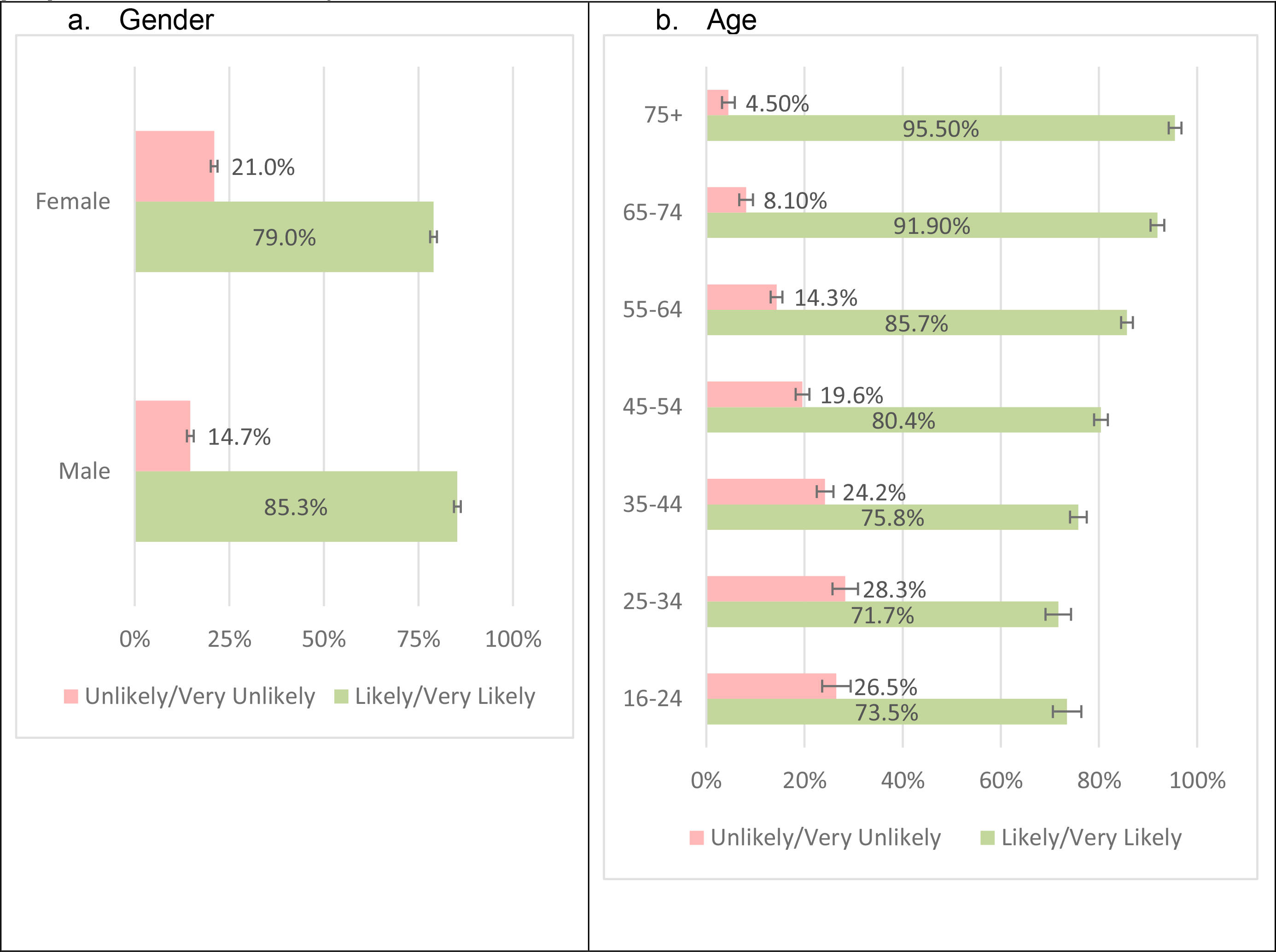
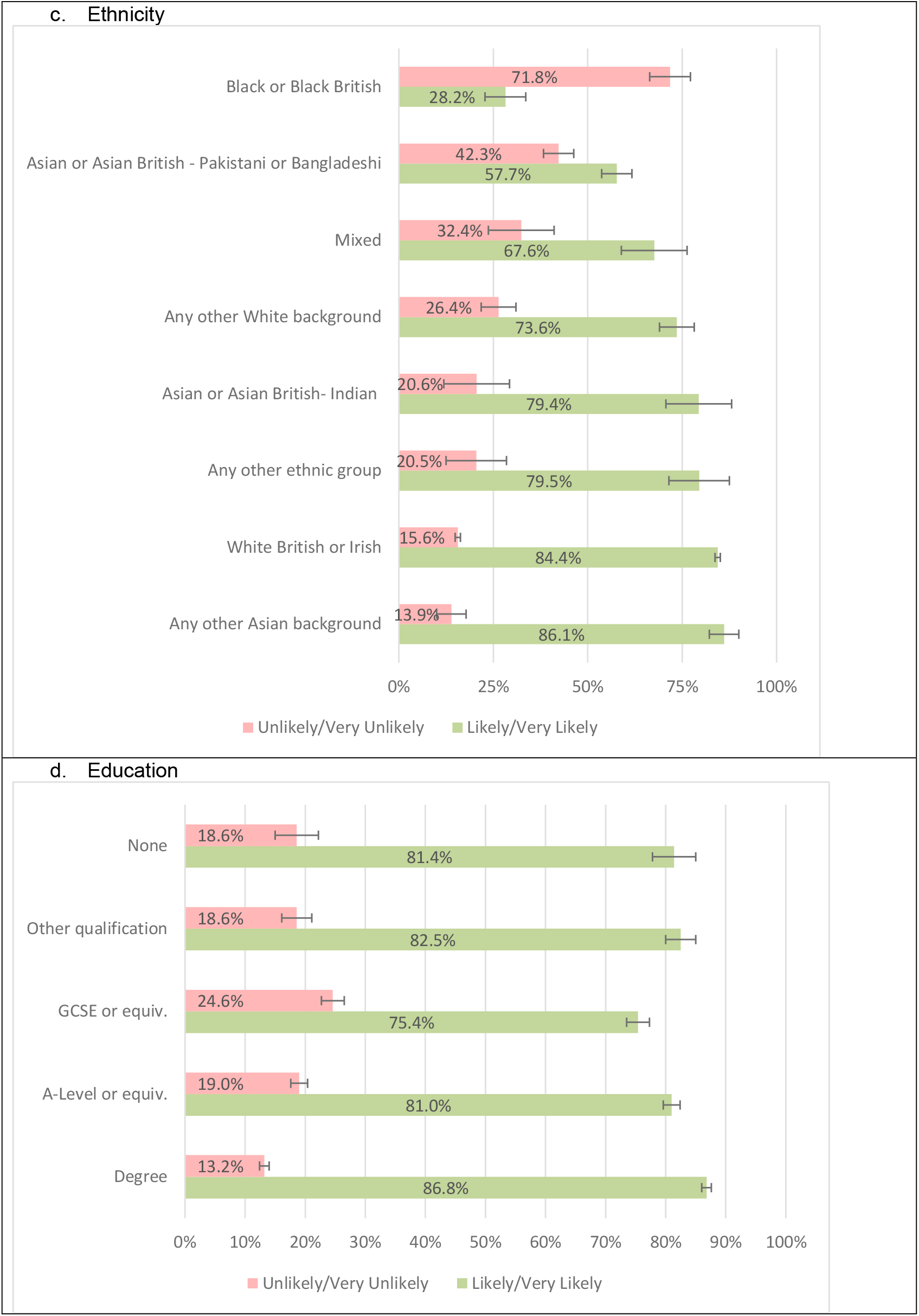
Proportions of vaccine hesitancy and willingness to be vaccinated (weighted proportions with 95% CI) Footnote: Question asked was “Imagine that a vaccine against COVID-19 was available for anyone who wanted it. How likely or unlikely would you be to take the vaccine?”

**Figure 3.**
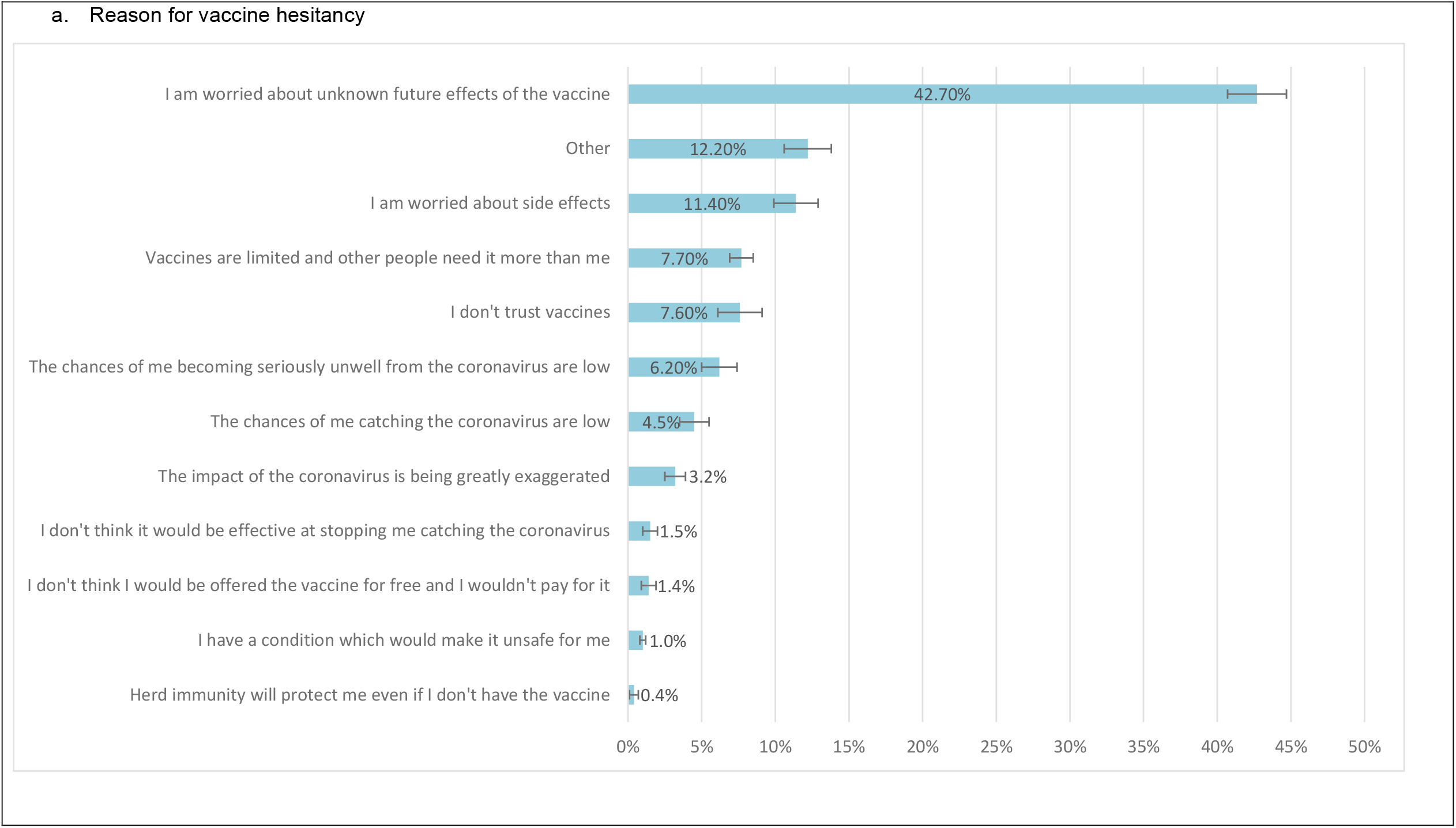
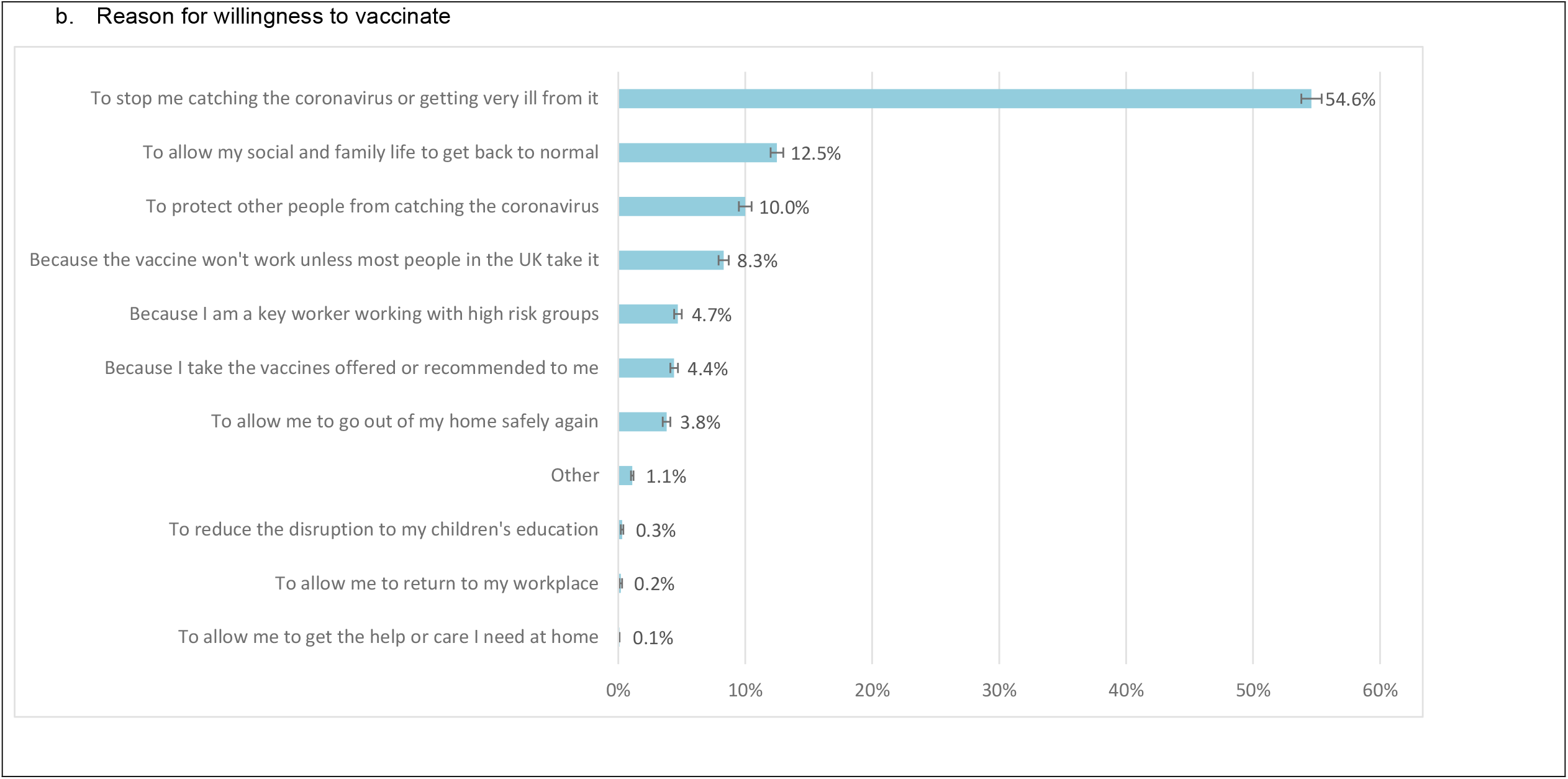
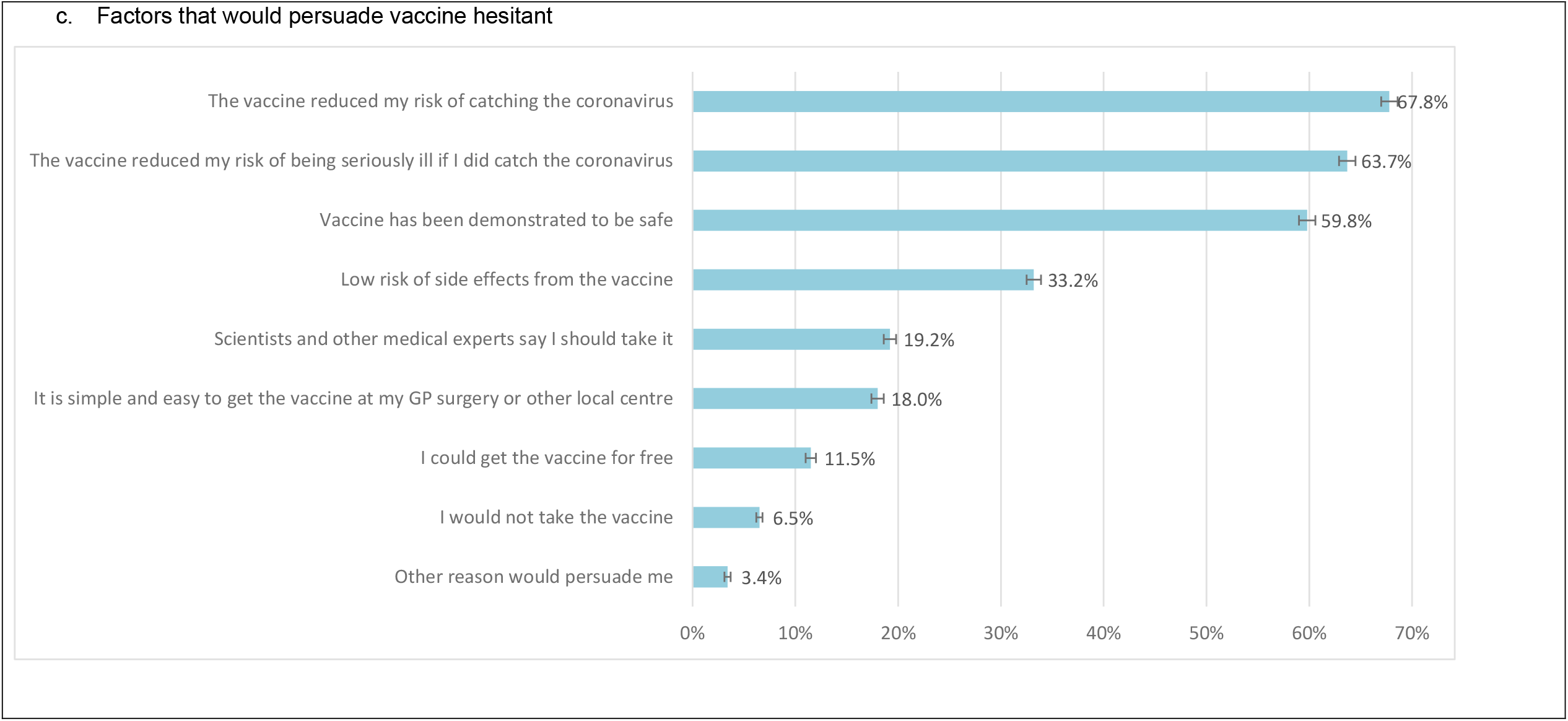
Reasons for vaccine hesitancy, willingness to take vaccines and factors that would persuade people to take a vaccine (weighted proportions with 95% CI bars) Footnote: Question for 3a “What is the main reason you would not take the vaccine?”; 3b *“What would be your main reason for taking the vaccine?”*; and 3c “*Which three of these things would most increase the chances of you choosing to get vaccinated?*”

### Covariates

Covariates are all taken from the longitudinal mainstage data files. We adjusted for age, coded as 16-24, 25-34, 35-44, 45-54, 55-64, 65-74, 75+, and gender coded as male/female. Ethnicity was self-reported and coded as White British or Irish; Other White; three Asian and Asian British groups: Indian, Pakistani/Bangladeshi, or Other Asian (includes participants of Chinese ethnicity); Black or Black British; and Other. The ethnicity groupings were chosen to allow analysis of as detailed ethnic groupings as possible, subject to having an adequate sample size for meaningful analysis in each category. Country of birth (UK/Not UK) and UK country of residence (England/Scotland/Wales/Northern Ireland) were also investigated. Education level was based on the highest qualification reported in the most recent wave (data collected in 2019) and coded as Degree & Other Higher Degree, A-Level or equivalent, GCSE or equivalent, other qualification and none. NHS Shielding category (Yes/No) was ascertained from previous COVID-19 survey waves on the basis of self-report (“*Have you received a letter, text or email from the NHS or Chief Medical Officer saying that you have been identified as someone at risk of severe illness if you catch coronavirus, because you have an underlying disease or health condition*?”).

### Statistical Analysis

Descriptive statistics (percentages and 95% confidence intervals (CI)) for the outcomes were calculated using cross-sectional weights to make the sample representative of the UK community dwelling population. Weights were calculated for each COVID-web survey based on differential non-response from wave 9. Predictors included in the weights were basic demographic factors, household composition, previous survey outcomes, COVID-19 survey paradata, such as the number of reminders issued, economic and health variables. The final weights are calculated as the inverse of the response propensity.^12^ Weights are not calculated for the 1,974 participants who did not take part at Wave 9, and hence they are excluded from the weighted analyses, hence the weighted sample is 9,981.

Logistic regression was used because the dependent variable was binary. Three models were considered: model 0 - univariate (i.e.no adjustment) models for each covariate, model 1 – each covariate adjusted for age and gender, model 2 – all covariates mutually adjusted.

Data were analysed using SPSS version 25 using the complex samples method to take account of the clustered and stratified sample. Missing data were excluded from the analyses using listwise deletion so each model may contain different numbers of participants.

## RESULTS

### Sample Statistics

12,035 participants completed the Covid-19 wave 6 survey online and the weighted sample is 9,981 (Figure 1). However, in the weighted sample 56 participants did not answer the vaccine hesitancy question and 591 participants had missing data on at least one covariate, and hence the weighted analytical sample for the full multivariable models is 9390, although in other models it varies based on the covariates included. The weighted study sample is described in Table 1.

**Table 1.** Description of the study population.

|  | Weighted n (%) |
| --- | --- |
| **Likelihood of taking up a coronavirus vaccination** |  |
| Very likely | 5306 (53.2%) |
| Likely | 2832 (28.4%) |
| Unlikely | 1131 (11.3%) |
| Very unlikely | 656 (6.6%) |
| Missing (including refusal and don't know) | 56 (0.5%) |
| **Gender** |  |
| Male | 4666 (46.8%) |
| Female | 5290 (53.0%) |
| Prefer not to say | 25 (0.3%) |
| **Age** |  |
| 16-24 | 920 (9.2%) |
| 25-34 | 1382 (13.8%) |
| 35-44 | 1542 (15.5%) |
| 45-54 | 1784 (17.9%) |
| 55-64 | 1938 (19.4%) |
| 65-74 | 1532 (15.3%) |
| 75+ | 882 (8.8%) |
| **Ethnicity** |  |
| White British or Irish | 8713 (87.3%) |
| Other white background | 269 (2.7%) |
| Mixed | 168 (1.7%) |
| Asian or Asian British - Indian | 176 (1.8%) |
| Asian or Asian British - Pakistani/Bangladeshi | 198 (2.0%) |
| Asian or Asian British - any other group | 106 (1.1%) |
| Black or Black British | 190 (1.9%) |
| Other Ethnic Group | 59 (0.6%) |
| Missing | 102 (1.0%) |
| **Born in UK** |  |
| Born in UK | 8991 (90.1%) |
| Not Born in UK | 824 (8.3%) |
| Missing | 166 (1.7%) |
| **UK Country** |  |
| England | 8424 (84.4%) |
| Wales | 507 (5.1%) |
| Scotland | 775 (7.8%) |
| Northern Ireland | 275 (2.8%) |
| **Highest Education Level** |  |
| Degree & Other Degree | 4086 (40.9%) |
| A Level or equivalent | 2202 (22.1%) |
| GCSE or equivalent | 2010 (20.1%) |
| Other Qualification | 846 (8.5%) |
| No Qualification | 501 (5.0%) |
| Missing(no education data in 2019 datafile (wave 10/11)) | 336 (3.4%) |
| **NHS Shielding Category** |  |
| Yes | 721 (7.2%) |
| No | 9244 (92.6%) |
| Don't know | 16 (0.2%) |

Unweighted N= 11,955, weighted N=9,981

### Prevalence of vaccine hesitancy

Overall, intention to have the COVID-19 vaccine was high, with 53.5% of participants saying they were very likely and a further 28.5% saying they were likely and 18% being vaccine hesitant (reporting unlikely or very unlikely). However, there was marked variation in population subgroups (Figure 2). A higher proportion of female participants indicated vaccine hesitancy, 21.0% compared to 14.7% of male participants. Younger age groups were also more vaccine hesitant with 28.3% of younger adults aged 25-34 vaccine hesitant compared to only 14.3% in the 55-64 age group, 8.1% in the 65-74 age group and 4.5% in the 75+ age group.

Black or black British were the ethnic group with the highest rate of vaccine hesitancy at 71.8%. Pakistani/Bangladeshi groups were the next most hesitant ethnic group with 42.3% vaccine hesitant, followed by those of Mixed ethnicity (32.4%). The ethnic groups with the highest intention to vaccinate were the White British and Irish groups (84.8% being likely/very likely to take a vaccine) and the any other Asian background group (86.1%; this group includes participants of Chinese ethnicity).

Vaccine hesitancy varied with education level. Vaccine hesitancy was lowest in those with degrees (13.2%) and highest in those with GCSE level education (24.6%).

### Reasons for vaccine hesitancy

The main reasons for vaccine hesitancy were concerns over future unknown effects of a vaccine, with 42.7% citing this as their main reason (Figure 3). The main reasons for being willing to take up a vaccine were to avoid catching COVID-19 or becoming ill from the disease (54.6%) and to allow social and family life to get back to normal (12.5%).

For those particular groups identified as vaccine hesitant, the main reasons given by Black/Black British participants for not taking the vaccine were a lack of trust in vaccines (29%) and worries about unknown future effects (30%). Pakistani/Bangladeshi participants cited concerns about side effects (36%) and unknown future effects (35%) as their main reasons for hesitancy. Women were more likely than men to say that their main reason for vaccine hesitancy was concern about side effects and to say they do not trust vaccines.

67.8% of participants reported that knowing a vaccine reduced their risk of catching COVID-19 was a factor that would increase their chance of taking a vaccine (Figure 3). Other factors commonly identified by participants as increasing their chances of taking a vaccine were if it reduced the risk of being seriously ill if they did catch COVID-19 (63.7%) and a vaccine being demonstrated to be safe (59.8%).

When asked what would most convince participants to take the vaccine, 43.2% of Black/Black British maintained that they would not take it, while a further 44.7% reported that they would if the vaccine was demonstrated to be safe. Pakistani/Bangladeshi participants reported that they may be persuaded if the vaccine reduced their risk of catching COVID-19 (65.2%) and/or if it was demonstrated to be safe (64.6%).

Females had higher odds of vaccine hesitancy than males (OR 1.55, 95%CI:1.31, 1.85) and this was still present in the mutually adjusted model (OR 1.68, 95% CI:1.42, 1.99) (Table 2). The risk of being vaccine hesitant was inversely related to age with younger age groups having higher odds of vaccine hesitancy; the odds in the 16-24 year old category were 1.48 (95% CI:1.06, 2.08) compared to those aged 45-54 years. However, once adjusting for other covariates this was reduced to 1.29 (95% CI: 0.91, 1.83). The vaccine hesitancy risk was higher in the 25-34 (OR 1.64, 95% CI: 1.19, 2.26) and 35-44 (OR 1.42, 95% CI:1.09, 1.85) than 45-54 year olds, and this was present in all models. Conversely those in older age groups were less likely to be vaccine hesitant across all models, 55-64 (OR 0.60, (95% CI:0.47, 0.77), 65-74 (OR 0.33, 95% CI:0.23, 0.45) and 75+ (OR 0.17, 95% CI:0.08, 0.34). For those participants who had received a letter about shielding, the odds of vaccine hesitancy were not different to those who are not shielding but were imprecisely estimated, and this was true across all models (OR 1.08, 95%CI:0.70, 1.66).

**Table 2.**
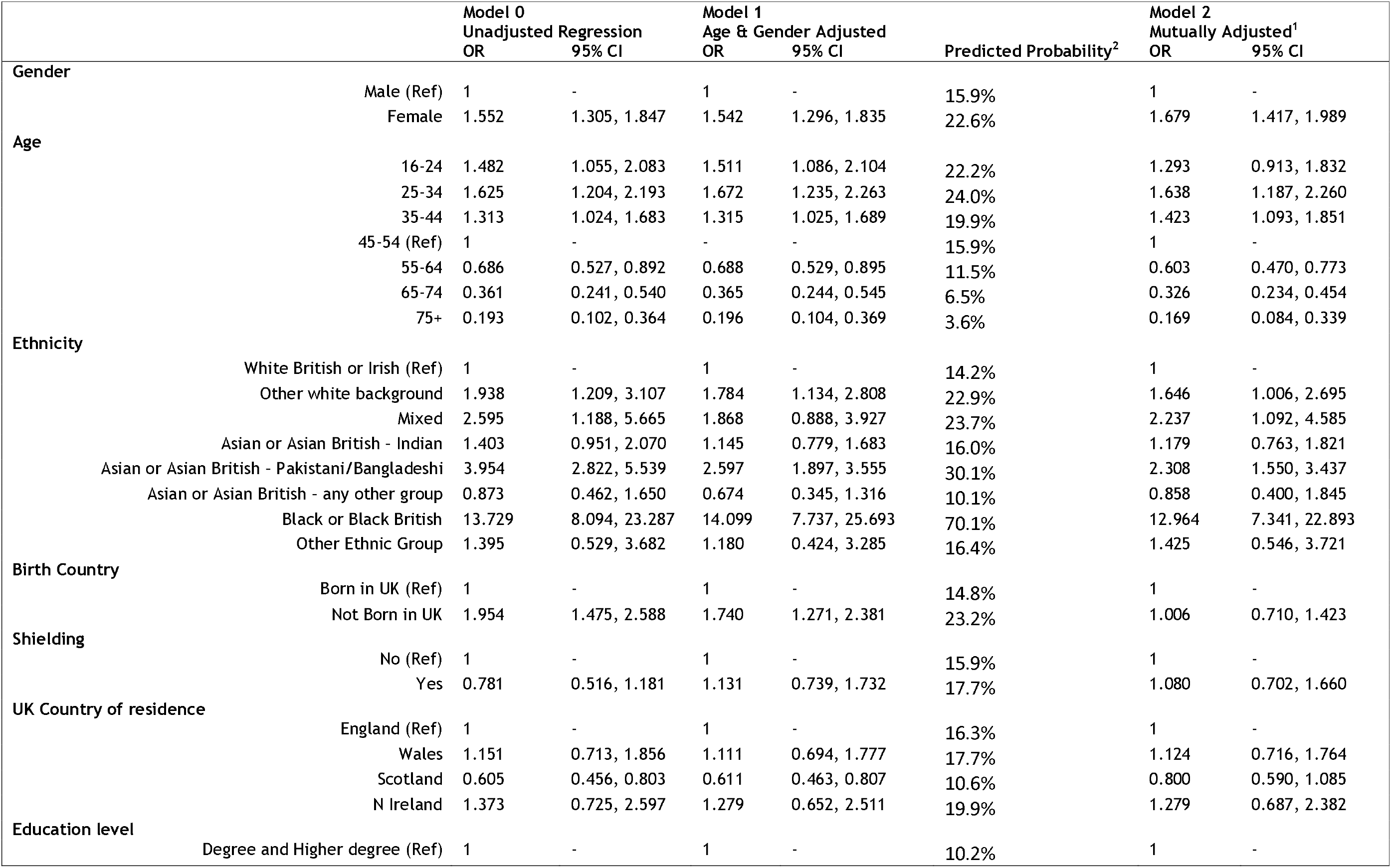
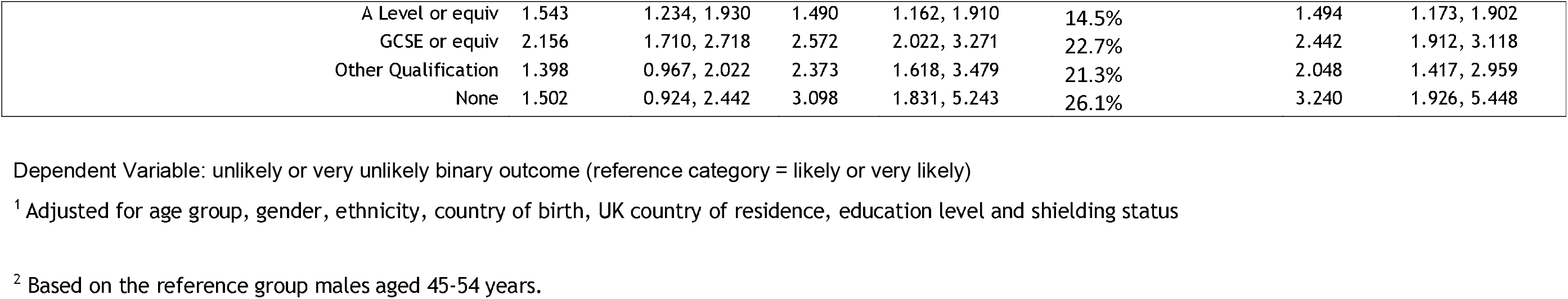
Logistic regression models for vaccine hesitancy.

Higher vaccine hesitancy was seen in most minority ethnic groups compared to the White British group. The highest odds were seen in the Black and Black British group (OR 12.96, 95% CI:7.34, 22.89) and the Pakistani and Bangladeshi groups (OR 2.31, 95% CI:1.55, 3.44), and adjustment for covariates made relatively little difference to these associations. Participants who were not born in the UK did not have greater odds of vaccine hesitancy (OR 1.01 95%CI: 0.71, 1.42) in the mutually adjusted model. In the mutually adjusted model, there were no substantive differences in vaccine hesitancy by country of residence.

The risk of vaccine hesitancy was inversely related to education in the mutually adjusted model. Compared to those with degree-level education there were raised odds of vaccine hesitancy among those with GCSE or equivalent qualifications (OR 2.05, 95% CI:1.42, 2.96), A-Level or equivalent (OR 1.49, 95% CI:1.17, 1.90), and no educational qualifications (OR 3.24, 95% CI:1.93, 5.45), indicating a socioeconomic gradient in vaccine hesitancy.

## DISCUSSION

The vast majority (82%) of UK adults are willing to take up a COVID-19 vaccine if offered but marked differences exist across population subgroups. Older age and being male are strongest drivers of the risk of COVID-19 death, they are less likely to be vaccine hesitant, suggesting the vaccination programme could yield large health benefits within the UK. Very large differences in vaccine hesitancy exist by ethnicity, with Black/Black British and Pakistani/Bangladeshi ethnic groups being most hesitant. However, not all minority ethnic groups had higher vaccine hesitancy, highlighting the importance of understanding heterogeneity between minority ethnic groups. Overall, the main reasons given for vaccine hesitancy were fears around side effects and future adverse effects of a COVID-19 vaccine. The main reasons for intended vaccine uptake relate to the avoidance of catching the virus or becoming very ill from it, but also to allow social and family life to return to normal. Vaccine efficacy and safety were identified as factors that would encourage vaccine uptake.

Our study has several strengths. To our knowledge, it is the first large representative study in the UK to survey participants on likely vaccine uptake or hesitancy and the reasons why a COVID-19 vaccine would be accepted or refused. It also did so at a time when new vaccine development and vaccine efficacy was highly reported in the media (end November 2020, but just before the first vaccine was approved on 2^nd^ December). The four countries of the UK are now rolling out a vaccination programme commencing with those most at risk of mortality from COVID-19. The findings reported here, provide evidence on the groups who need targeting and arguments that may be most persuasive for them. However, there are limitations which should be noted. The survey is web-based so non-participation may have introduced bias into the results. However, the results were weighted to account for non-response and attrition. Small numbers did not allow for detailed analysis of some ethnic groups. Additionally, we did not ask about the different types of vaccinations being developed and whether this would have any bearing on vaccine hesitancy. While we highlight associations between vaccine hesitancy and range of socio-demographic factors, the purpose of this analysis was descriptive. Willingness to be vaccinated is influenced by public health and other communications, as well as a broader range of social factors. These associations should therefore not be interpreted as immutable effects but rather guide vaccination planning.

There have been some other smaller UK studies of COVID-19 vaccine hesitancy which have not been based on representative samples. These studies indicated that 14% of participants were unwilling to receive a COVID-19 vaccine with a further 22% being unsure as to whether they would take this, with only 64% saying they would take a COVID-19 vaccine.^15^ A Scottish survey found uptake figures to be slightly higher at between 78-81%.^16^ A poll by Ipsos MORI in late October 2020 found 67% of the UK public said they were very (42%) or fairly (25%) likely to take a COVID-19 vaccine.^17^ Our study suggests slightly higher vaccine uptake in the general UK population of 82% with vaccine hesitancy at 18%.

Given the age related focus of the Phase One vaccination roll out in the UK ^18^, our study suggests that uptake is likely to be high in the target groups. However, the finding of greater vaccine hesitancy amongst some, but not all, ethnic minority groups is concerning and aligns with emerging evidence from other countries. This is not inevitable – studies focused on intention to receive vaccines prior to this pandemic have not consistently found greater levels of vaccine hesitancy among ethnic minority groups.^19,20^ Nevertheless, it needs to be a key focus of the design of vaccination programmes, given the higher prevalence of COVID-19 among ethnic groups. Our study also found that those who had lower education levels were more likely to be vaccine hesitant suggesting that there similarly needs to be added focus on increasing vaccine uptake with those experiencing socioeconomic disadvantage who are at more risk of COVID-19.^21^

Vaccine hesitancy rates vary by country and population subgroup. However, international evidence across several studies suggests that approximately 25% of the general population are hesitant about accepting a COVID-19 vaccine.^22-27^ A systematic review of studies on willingness to be vaccinated suggests 60% of people intend to be vaccinated, indicating the UK may be better placed to utilise vaccination to address the pandemic.^28^ In some countries, such as Italy, previous vaccination rates suggest uptake may be too low to stop the spread of COVID-19.^29^ Our study suggests that this is not the case in the UK and if everyone who has said they are likely or very likely to take up the vaccination if offered, actually get vaccinated, coverage in the UK could be high enough to achieve herd immunity. Existing research for vaccine hesitancy suggests that key drivers for uptake were perceived efficacy of a vaccine, concern over negative adverse effects and safety of a vaccine.^15,25,30^ Our study also found fears over adverse effects and safety to be key reasons for vaccine hesitancy, especially future negative effects. However, our study also adds that knowing that a vaccine is effective in reducing the spread of COVID-19 could increase uptake.

Our study has important practical implications for public health policy. There are identifiable subgroups of the UK population who are more likely to be vaccine hesitant. Vaccine hesitancy is a complex problem ^31^ and as such a range of practical steps need to be undertaken to increase uptake. Firstly, the subgroups we have identified as being vaccine hesitant should be included in the planning and development of any engagement programmes. There is the potential to widen health inequalities without deliberate efforts to engage those groups who are most likely to be affected by COVID-19 and least likely to take up a vaccine. Initiatives to improve uptake in Black ethnic groups within the UK should be an urgent priority – for example, by working in close partnership with communities and making use of community champions.^32^ While universal and targeted educational interventions are necessary to enable the public to understand the importance of vaccination and are ethically and politically acceptable, they are not enough to modify behaviour or increase confidence.^33^ Full endorsement from regulatory bodies is likely to increase confidence^34^, but efforts to combat misinformation, especially around vaccine safety, may be warranted. The rise in vaccine hesitancy as a result of misinformation about safety coincides with the rise in social media, a growing platform for the anti-vaccination movement.[22,23] A concerted effort to engage with younger adults both online and through traditional communication channels will be needed if confidence in a vaccine is to be achieved and vaccine uptake is to improve in this group, subject to them being included in future vaccination rollout.

Further detailed qualitative research should investigate the reasons for vaccine hesitancy with the subgroups identified as highly hesitant and approaches to overcoming them. While compulsory vaccination is unlikely in the UK, we have not asked about the acceptability of mandated vaccination for certain situations, e.g. immunisation passports or restrictions based on vaccination status. Further research would be required to understand whether a form of mandating vaccination would be acceptable to the UK population, for example only allowing those vaccinated to visit care homes or travel restrictions based on vaccination status. As vaccination programmes continue to be implemented, ongoing monitoring of uptake and vaccination attitudes are needed.

## Data Availability

UK Household Longitudinal Study data are available from the UK Data Archive.

## FOOTNOTES

### Contributors

MJB and SVK conceived, designed the study and contributed equally to this paper. ER drafted the manuscript. KSR implemented the statistical analyses. ER & KSR contributed equally to this paper. MB led on question design for the vaccine questions. JM calculated weights. All authors critically revised the article, contributed to data interpretation, and finalised and approved the manuscript.

